# Investigating SARS-CoV-2 breakthrough infections per variant and vaccine type

**DOI:** 10.1101/2021.11.22.21266676

**Authors:** Jozef Dingemans, Brian M.J.W. van der Veer, Koen M.F. Gorgels, Volker Hackert, Audrey Y.J. Hensels, Casper D.J. den Heijer, Christian J.P.A Hoebe, Paul H.M. Savelkoul, Lieke B. van Alphen

## Abstract

Breakthrough SARS-CoV-2 infections have been reported in fully vaccinated individuals, in spite of the high efficacy of the currently available vaccines, proven in trials and real-world studies. Several variants of concern (VOC) have been proffered to be associated with breakthrough infections following immunization. In this study, we investigated 378 breakthrough infections recorded between January and July 2021 and compared the distribution of SARS-CoV-2 genotypes identified in 225 fully vaccinated individuals to the frequency of circulating community lineages in the region of South Limburg (The Netherlands) in a week-by-week comparison. Although the proportion of breakthrough infections was relatively low and stable when the Alpha variant was predominant, the rapid emergence of the Delta variant lead to a strong increase in breakthrough infections, with a higher relative proportion of individuals vaccinated with Oxford-AstraZeneca or J&J/Janssen being infected compared to those immunized with mRNA-based vaccines. A significant difference in median age was observed when comparing fully vaccinated individuals with severe symptoms (83 years) to asymptomatic cases (46.5 years) or individuals with mild-to-moderate symptoms (42 years). There was no association between SARS-CoV-2 genotype or vaccine type and disease symptoms. Interestingly, symptomatic individuals harbored significantly higher SARS-CoV-2 loads than asymptomatic vaccinated individuals and breakthrough infections caused by the Delta variant are associated with increased viral loads compared to those caused by the Alpha variant. Altogether, these results indicate that the emergence of the Delta variant might have lowered the efficiency of particular vaccine types to prevent SARS-CoV-2 infections and that, although rare, the elderly are particularly at risk of becoming severely infected as the consequence of a breakthrough infection.

## Introduction

Since its discovery in December 2019^1^, SARS-CoV-2 has infected more than 250 million people and caused about 5 million deaths^2^. Although SARS-CoV-2 has a limited mutation rate of about 2 mutations per month^3^, which can be attributed to its 3’ to 5’ exonuclease proofreading activity^4^, its rapid expansion across the globe has led to the emergence of numerous variants^5,6^. Whereas the majority of variants initially harbored mutations outside the gene encoding the spike protein, a number of variants have emerged with mutations in the spike protein that affect infectivity or immune evasion. For example, the mutation leading to the D614G substitution in the spike protein emerged soon after SARS-CoV-2 spread to Europe at the beginning of the first wave^7^. This mutation greatly affected the fitness of SARS-CoV-2 by stabilizing the open conformation of the spike protein, enhancing viral infectivity and leading to dominance of this variant across the globe^7-9^. More recently, a number of variants of concern (VOC) have emerged that harbor mutations in the receptor binding domain (RBD) of the spike protein, enhancing the affinity for the human angiotensin-converting enzyme 2 (ACE2) receptor^10^ or leading to evasion of the immune system^11^. In November 2020, the B.1.1.7 variant was first reported as it quickly gained dominance in the UK, imposing a great burden on the healthcare system due to increased infectivity and slightly heightened mortality^12-14^. Both the N501Y and V69/H70del mutations in the spike protein of B.1.1.7 have been shown to enhance its infectivity^10,14,15^, while the Y144del mutation was found to be in the antigenic supersite^16^. Although the B.1.1.7 variant has been associated with enhanced infectivity, it has been shown that sera from previously infected and vaccinated individuals as well as monoclonal antibodies can efficiently neutralize this variant *in vitro*^17-19^. Nevertheless, two other VOC (B.1.351 and P.1) have been identified that exhibit significantly reduced neutralization by monoclonal antibodies or sera from recovered or vaccinated individuals^6,11,17-23^. The B.1.351 variant that was first reported in South Africa and the P.1 variant that was identified in Manaus, Brazil, share the E484K and K417N mutations in the RBD that are associated with immune evasion as well as the N501Y mutation, which is also present in B.1.1.7 and presumably enhances infectivity. Finally, a fourth VOC (B.1.617.2) has recently been identified in India, harboring the L452R and T478K mutations in RBD of the spike protein, showing moderately reduced neutralization and increased transmissibility compared to B.1.1.7^20,24-26^.

Although many studies have assessed the potential of these variants to evade the immune system *in vitro*, only a limited number of studies have looked into the distribution of variants that can cause infections in fully vaccinated individuals. Monitoring these so-called post-vaccination ‘breakthrough variants’ is imperative since they may signal widespread reduced vaccine efficacy early on. In this study, we have compared the occurrence of variants causing infections in 378 fully vaccinated individuals to the prevalence of variants that were identified by regional surveillance of SARS-CoV-2 in the South Limburg region of the Netherlands from January to July 2021.

## Methods

### Case definition

In this study, cases were defined as patients who were fully vaccinated against SARS-CoV-2 (≥14 days post-2nd dose of Pfizer-BioNTech, Moderna or Oxford-AstraZeneca vaccine or ≥14 days post 1 dose of Johnson&Johnson vaccine) who had onset of COVID-19 related symptoms and subsequently tested positive by real-time polymerase chain reaction (RT-PCR) or antigen test for SARS-COV-2.

Cases were defined as symptomatic if they reported COVID-19 related symptoms, including common cold symptoms (nasal cold, runny nose, sneezing or sore throat), cough, elevated temperature or fever (temperature > 38°C), loss of taste or smell, diarrhea, nausea, fatigue, headache and generalized pain. Cases were defined as asymptomatic if they reported no COVID-19 related symptoms at the time of their positive test and developed no symptoms during the 7 day follow-up.

Symptomatic cases were further subdivided in mild-moderate and severe cases. All cases that were hospitalized or had a fatal outcome were classified as severe. All other symptomatic cases were classified as mild-moderate.

All post-vaccination breakthrough infections described were investigated by the Public Health Service South Limburg, the Netherlands. Cases or relevant staff members were contacted in order to gain missing data.

### Sampling and RT-PCR assay

Trained personnel collected combined nasopharyngeal/oropharyngeal swabs in viral transport medium. Samples were sent either to the Medical Microbiological Laboratory of Maastricht University Medical Centre (MUMC+) (workflow 1) or Synlab Belgium (workflow 2) for laboratory confirmation of SARS-CoV-2 via RT-PCR assay.

#### Workflow 1: RT-PCR analysis at the MUMC+

Samples were collected and transported in viral transport medium (Mediaproducts, The Netherlands). For RNA extraction, 900 µl of clinical sample was mixed with 900 µl of Chemagic Viral Lysis Buffer (Perkin-Elmer) and RNA was extracted from samples using the Chemagic Viral DNA/RNA 300 Kit H96 (Perkin-Elmer) on the Chemagic 360 system (Perkin-Elmer). A multiplex RT-PCR was performed using the N1-gene and E-gene as targets, including the immediate early gene of mouse cytomegalovirus as an internal control (Table S1). cDNA synthesis and PCR amplification were combined using the TaqPath™ 1-Step RT-qPCR Master Mix, CG (Applied Biosystems, US). Thermal cycling was performed using the Quantstudio 5 Real-Time PCR System (Applied Biosystems, US). Oligonucleotides were synthesised and provided by Biolegio (Netherlands) (Table S1). Only the CT-value obtained for the N1 target was reported.

#### Workflow 2: RT-PCR analysis at Synlab Belgium

Alternatively, RT-PCR analysis was performed at Synlab Belgium using the ORF1ab, S-gene, and N-gene as targets. CT-values were reported for all three targets.

### Sequencing of SARS-CoV-2-positive samples

Sequencing was performed using the PCR tiling of SARS-CoV-2 virus with Native Barcoding Expansion 96 (EXP-NBD196) protocol (Version: PTCN_9103_v109_revH_13Jul2020) of Oxford Nanopore technologies, with minor modifications and using the primers previously published by Oude Munnink et al. ^27^ Briefly, the only modifications were extending the barcode and adaptor ligation steps up to 60 min and loading 48 samples per flow cell. Bioinformatic analysis was performed using an in-house developed pipeline MACOVID (https://github.com/MUMC-MEDMIC/MACOVID) that is based on Artic v1.1.3. Pangolin lineages were assigned using the Pangolin COVID-19 Lineage Assigner web application on https://pangolin.cog-uk.io/.

Consensus sequences with >3000 Ns were considered as low quality and therefore excluded for further analyses. However, consensus sequences <10000 Ns with a complete spike protein sequence were included.

### Mapping of non-lineage defining mutations on SARS-CoV-2 spike trimer model

The pdb file of the model 7D3F of the structure of the SARS-CoV-2 (Wuhan-Hu-1 strain) spike trimer in closed conformation as determined via cryo-electron microscopy by Xu *et al*^28^ (doi: 10.2210/pdb7DF3/pdb) was visualized using the UCSF Chimera protein viewer^29^. Non-lineage defining mutations were highlighted in red using the labeling tool in the Chimera protein viewer.

### Statistical analysis

All statistical analyses were performed using GraphPad Prism 9.0.0 software (GraphPad, La Jolla, CA, USA). A Mann-Whitney test was used to compare CT-values from breakthrough infections with the Alpha variant vs infections caused by the Delta variant. A Kruskal-Wallis test was performed to compare median age, time of positive test post 2^nd^ dose, and CT value between groups (symptoms or vaccine types). Dunn’s multiple comparison test was performed *post hoc* in case of a significant difference to identify which groups significantly differed. Fisher’s exact test was performed to investigate the relationship between SARS-CoV-2 genotype (Alpha or Delta) and symptoms (asymptomatic or symptomatic), while a Chi square test was performed to investigate the relationship between vaccine type and symptoms.

### Medical Ethical Approval

All data were retrieved from regular infectious disease control activities and were deidentified. The Medical Review Ethics Committee of the Maastricht UMC+ confirmed that the Medical Research Involving Human Subjects Act (WMO) does not apply to the above mentioned study and that an official approval of this study by the committee is not required (METC reference number 2021-2838).

## Results

### Description of the overall study population

In total, 378 breakthrough infections were recorded in the South Limburg region of the Netherlands until the end of week 28. This study population had a median age of 45 years, consisted for 63% of women and most individuals (62%) received 2 doses of the Pfizer-BioNTech vaccine (Table 1). The majority of cases were detected via community screening, while 8% of cases were related to care facilities (4% residents + 4% employees) and 14 cases (3.7%) were related to outbreaks (Table 1, Table S2). The majority of people (75%) experienced mild to moderate symptoms and there were 2 fatalities amongst the 7 severe cases (Table 1, Table S2).

**Table 1.**
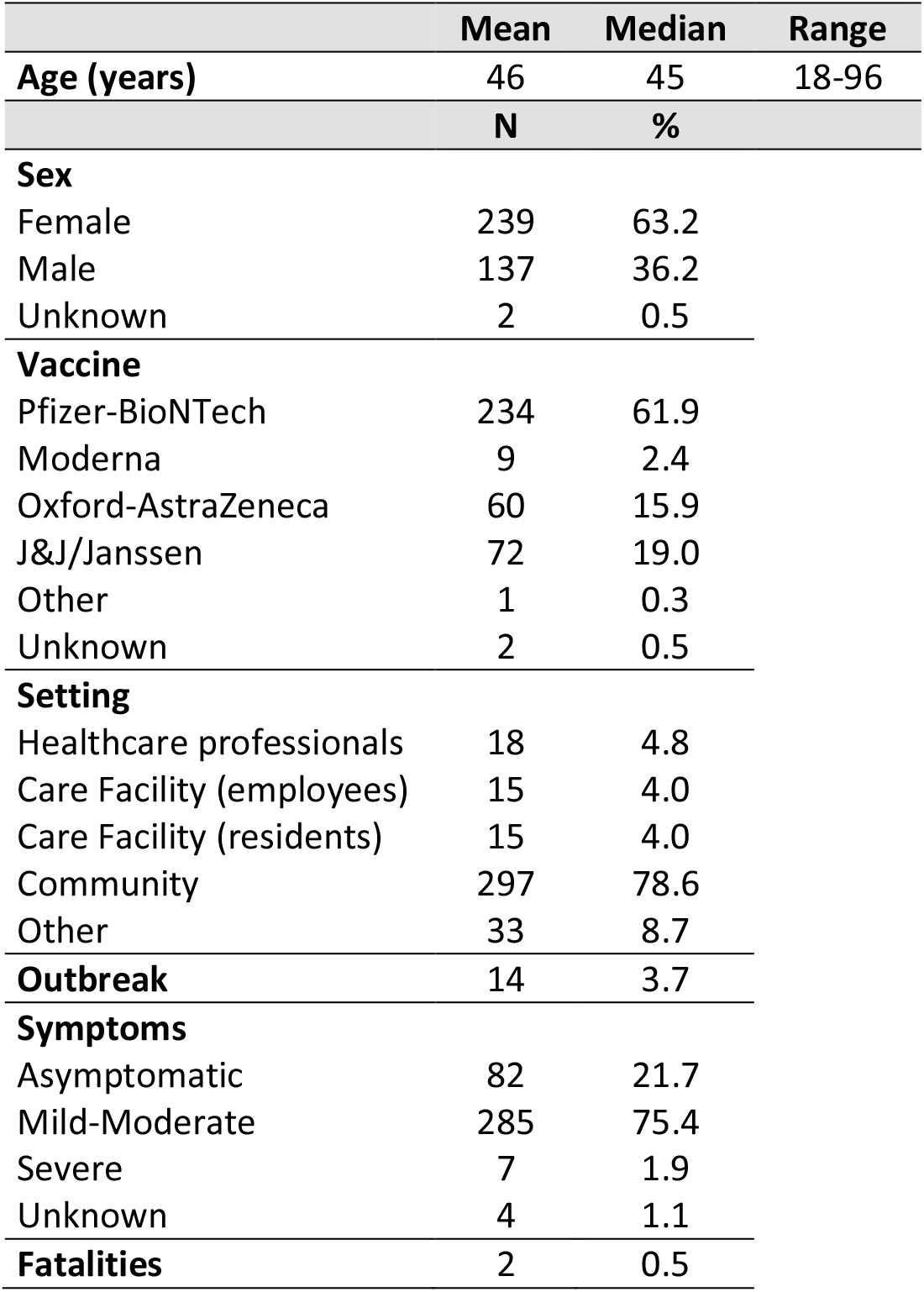
Characteristics of the study population.

|  | Mean | Median | Range |
| --- | --- | --- | --- |
| **Age (years)** | 46 | 45 | 18-96 |
|  | **N** | **%** |  |
| **Sex** |  |  |  |
| Female | 239 | 63.2 |  |
| Male | 137 | 36.2 |  |
| Unknown | 2 | 0.5 |  |
| **Vaccine** |  |  |  |
| Pfizer-BioNTech | 234 | 61.9 |  |
| Moderna | 9 | 2.4 |  |
| Oxford-AstraZeneca | 60 | 15.9 |  |
| J&J/Janssen | 72 | 19.0 |  |
| Other | 1 | 0.3 |  |
| Unknown | 2 | 0.5 |  |
| **Setting** |  |  |  |
| Healthcare professionals | 18 | 4.8 |  |
| Care Facility (employees) | 15 | 4.0 |  |
| Care Facility (residents) | 15 | 4.0 |  |
| Community | 297 | 78.6 |  |
| Other | 33 | 8.7 |  |
| **Outbreak** | 14 | 3.7 |  |
| **Symptoms** |  |  |  |
| Asymptomatic | 82 | 21.7 |  |
| Mild-Moderate | 285 | 75.4 |  |
| Severe | 7 | 1.9 |  |
| Unknown | 4 | 1.1 |  |
| **Fatalities** | 2 | 0.5 |  |

### Evolution of breakthrough cases in function of time and per vaccine type

The first individuals in the region of South Limburg received their 2^nd^ dose of the Pfizer-BioNTech vaccine in week 4 and the number of people who were fully vaccinated steadily increased starting from week 6 (Figure 1A). The majority of people received an mRNA-based vaccine (mostly Pfizer-BioNTech) and only starting from week 9, the first individuals were fully vaccinated with Oxford-AstraZeneca, while the first fully vaccinated individuals who received the J&J/Janssen vaccine started accumulating in week 21.

**Figure 1.**
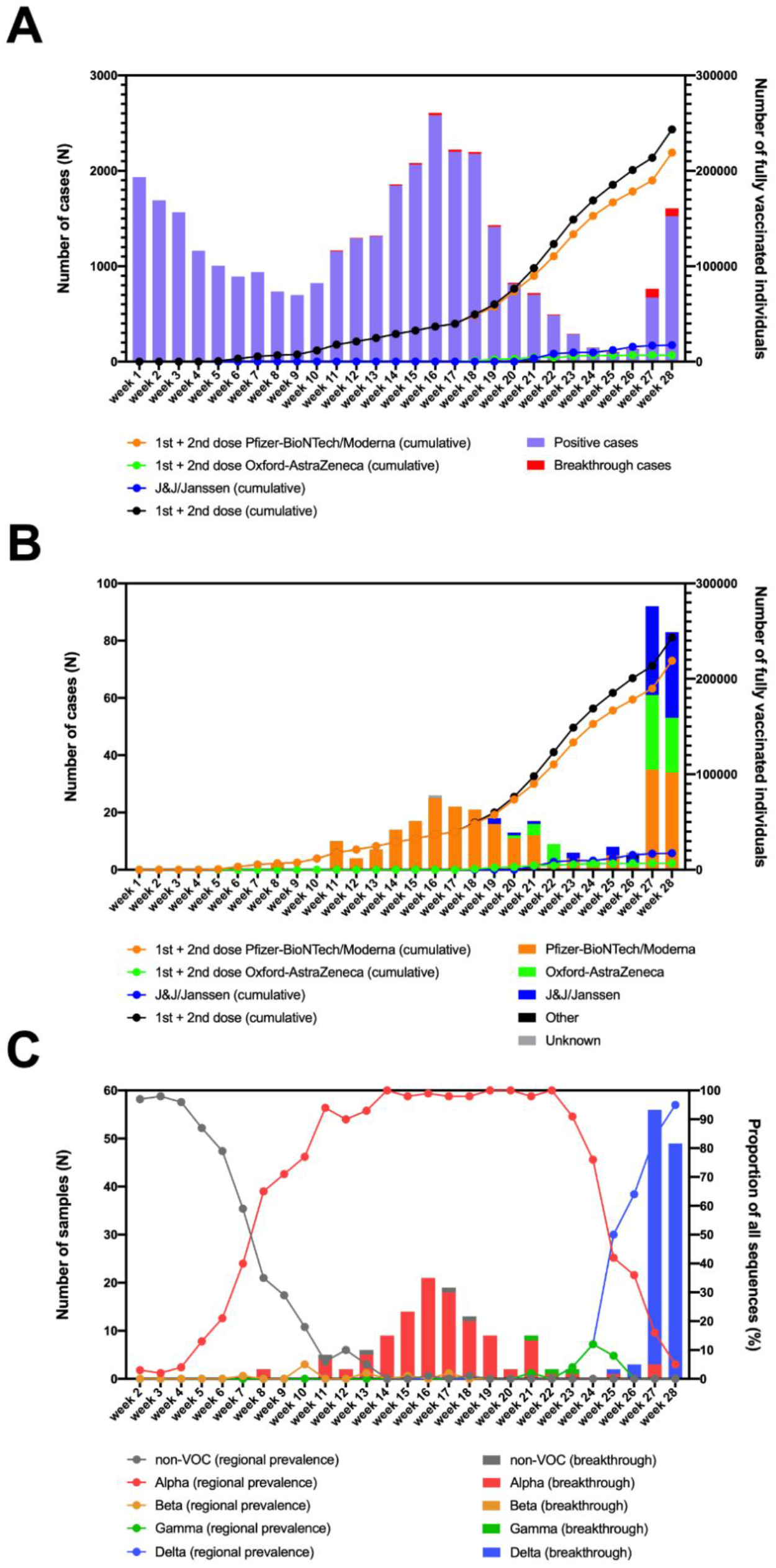
**A**. Overview of the number of fully vaccinated individuals and breakthrough infections in function of time in the region of South Limburg. **B**. Breakthrough infections per vaccine in function of time. **C**. Evolution of the prevalence of Non-VOC and VOC lineages in the community (lines) as well as the distribution of variants causing breakthrough infections (bars) in function of time in the region of South Limburg. The numbers represented by bars are plotted on the left y-axis, while the numbers represented by dots/lines are plotted on the right y-axis.

The first breakthrough infections were reported in week 8 and continually occurred starting from week 11, with a frequency ranging between 0.3 to 13.7% relative to the total number of infections (Figure 1A). The number of breakthrough cases steeply increased in week 27, after most COVID-19 measures were relaxed in The Netherlands and comprised disproportionally higher numbers of individuals who received Oxford-AstraZeneca or J&J/Janssen compared to the mRNA-based vaccines Pfizer-BioNTech/Moderna (Figure 1B). This was reflected by a considerably lower number of breakthrough cases relative to the number of administered vaccines for Pfizer-BioNTech/Moderna (0.14%) compared to Oxford-AstraZeneca (0.89%) or J&J/Janssen (0.46%) (Table 2).

**Table 2.**
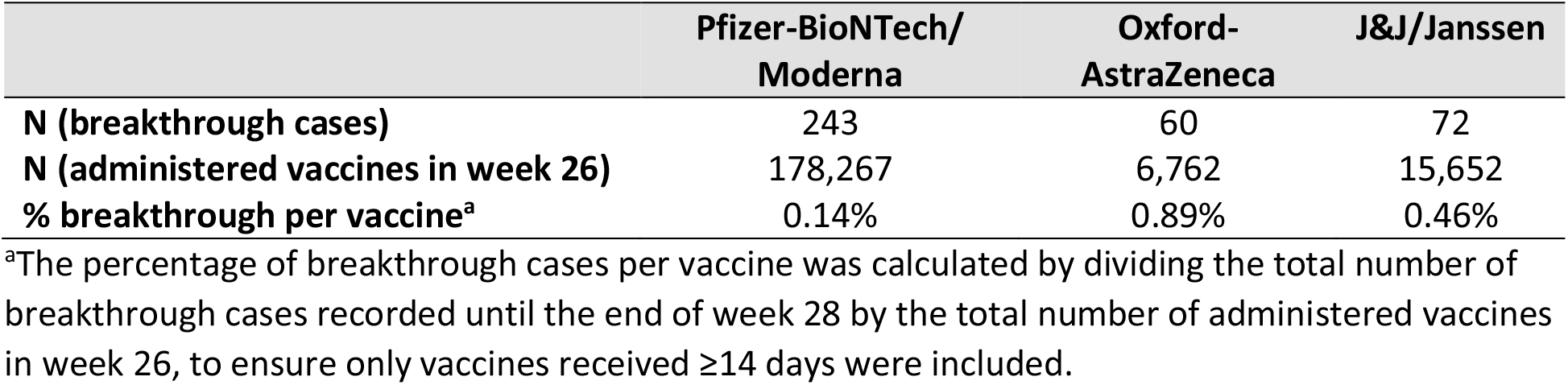
Overview of breakthrough cases and administered vaccines per vaccine type.

|  | Pfizer-BioNTech/<br>Moderna | Oxford-AstraZeneca | J&J/Janssen |
| --- | --- | --- | --- |
| **N (breakthrough cases)** | 243 | 60 | 72 |
| **N (administered vaccines in week 26)** | 178,267 | 6,762 | 15,652 |
| **% breakthrough per vaccine<sup>a</sup>** | 0.14% | 0.89% | 0.46% |

<sup>a</sup>The percentage of breakthrough cases per vaccine was calculated by dividing the total number of breakthrough cases recorded until the end of week 28 by the total number of administered vaccines in week 26, to ensure only vaccines received $\geq 14$ days were included.

### The distribution of SARS-CoV-2 variants in fully vaccinated individuals generally coincides with their regional prevalence

From the 378 breakthrough cases that were identified from week 2 until week 28, 225 were successfully genotyped via whole-genome sequencing. The remaining samples had a Ct-value >32 (n=19), were unavailable for sequencing (n=83), or did not comply to quality requirements of sequencing (n=51).

At the same time, regional surveillance of SARS-CoV-2 variants was performed on a weekly base. The non-VOC genotypes, which mainly consisted of the B.1.177 and B.1.221 lineages that have been prominent in the region since the summer of 2020^30^, were responsible for more than 50% of SARS-CoV2 infections up to week 7, after which the Alpha variant became dominant (Figure 1B). From this point, the frequency of Alpha infections rose quickly, constituting >90% of SARS-CoV-2 cases in week 11. In week 8 the first two breakthrough cases were identified, belonging to the Alpha variant (Figure 1B). Starting from week 11, the great majority of SARS-CoV-2 breakthrough infections were caused by the Alpha variant, with the exception of an occasional Gamma or a non-VOC infection (Figure 1C). The first breakthrough infection caused by the Delta variant was detected in week 25, when this variant was responsible for 50% of cases in the region and became dominant. As COVID-19 measures were relaxed in The Netherlands by the end of week 25, the number of breakthrough cases peaked during week 27 and 28, of which the great majority was caused by the Delta variant that was also dominant at that moment as determined via regional surveillance. The proportion of breakthrough infections caused by the Delta variant seems to be slightly higher than its share in genomic surveillance (100% vs 64% in week 26, 95% vs 84% in week 27, and 100% vs 95% in week 28).

### Breakthrough cases infected with the Delta variant harbor a higher viral load compared to cases infected with the Alpha variant

To investigate whether infections caused by the Delta variant are associated with increased viral loads compared to infections with the Alpha variant in fully vaccinated individuals, we compared the CT values obtained for the N1 target as determined via a single workflow. A significantly lower CT value (median CT-value = 17) was observed for breakthrough infections caused by the Delta variant vs those caused by the Alpha variant (median CT-value = 22) (Figure 2).

**Figure 2.**
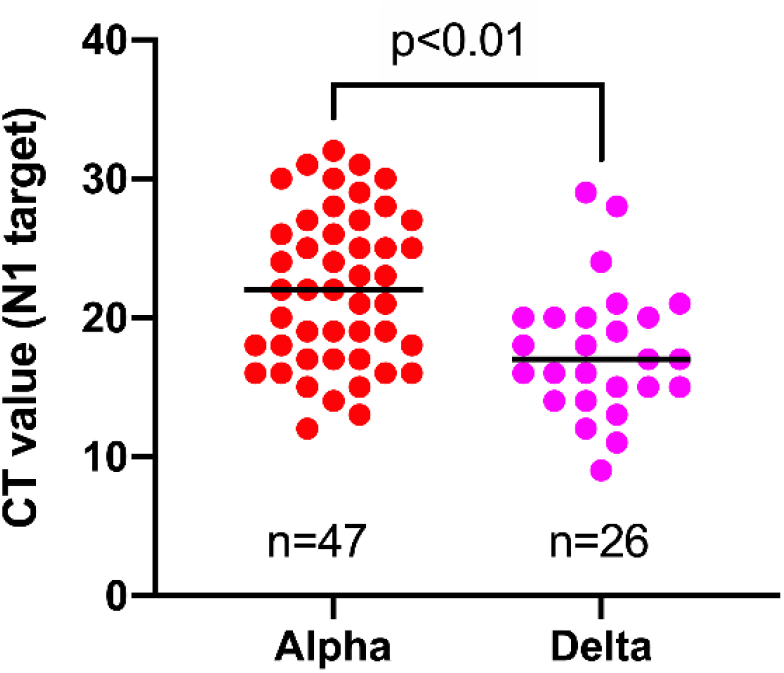
Comparison of CT values obtained by the same workflow for samples harboring Alpha versus Delta variant in breakthrough cases.

### Non-lineage defining mutations in the spike protein are mainly concentrated in the N-terminal domain amongst isolates causing breakthrough infections

Of the 225 isolates that were sequenced, 4 were non-variants of concern (non-VOC), 112 Alpha variant, 3 Gamma, and 106 Delta variant. In total, 46 non-lineage defining mutations were identified in the spike protein of the 225 isolates that were sequenced (Table 2), of which nearly half (19/46) were located in the N-terminal domain, while only a small minority (5/46) was found in the receptor binding domain (RBD) (Table 3 & Figure 3), despite both domains being of comparable size. Nine of the ten most frequent mutations (>1% frequency) occurred in the N-terminal domain (6/10) or in the C-terminal domain (3/10) and the two most observed mutations R21T (10.7%) and A222V (28.4%) were identified in at least 2 different variant backgrounds.

**Table 3.** Non-lineage-defining mutations in the spike protein found in this study.

| Mutation | N<sup>a</sup> | Frequency (%) | N (%) in non-VOC | N (%) in Alpha | N (%) in Gamma | N (%) in Delta |
| --- | --- | --- | --- | --- | --- | --- |
| L5F | 8 | 3.6% | 0 (0%) | 8 (7%) | 0 (0%) | 0 (0%) |
| S12F | 1 | 0.4% | 0 (0%) | 1 (1%) | 0 (0%) | 0 (0%) |
| L18F | 1 | 0.4% | 0 (0%) | 1 (1%) | NA<sup>b</sup> | 0 (0%) |
| R21T | 24 | 10.7% | 0 (0%) | 1 (1%) | 0 (0%) | 23 (22%) |
| T22I | 1 | 0.4% | 1 (25%) | 0 (0%) | 0 (0%) | 0 (0%) |
| T29I | 2 | 0.9% | 2 (50%) | 0 (0%) | 0 (0%) | 0 (0%) |
| T29A | 1 | 0.4% | 0 (0%) | 0 (0%) | 0 (0%) | 1 (1%) |
| A67V | 7 | 3.1% | 0 (0%) | 0 (0%) | 0 (0%) | 7 (7%) |
| T95I | 11 | 4.9% | 0 (0%) | 0 (0%) | 0 (0%) | 11 (10%) |
| K97Q | 1 | 0.4% | 0 (0%) | 0 (0%) | 0 (0%) | 1 (1%) |
| I128V | 1 | 0.4% | 0 (0%) | 1 (1%) | 0 (0%) | 0 (0%) |
| K147N | 3 | 1.3% | 0 (0%) | 3 (3%) | 0 (0%) | 0 (0%) |
| K150E | 1 | 0.4% | 0 (0%) | 1 (1%) | 0 (0%) | 0 (0%) |
| M153I | 1 | 0.4% | 0 (0%) | 1 (1%) | 0 (0%) | 0 (0%) |
| S221L | 1 | 0.4% | 0 (0%) | 0 (0%) | 0 (0%) | 1 (1%) |
| A222V | 64 | 28.4% | 1 (25%) | 0 (0%) | 0 (0%) | 63 (59%) |
| T250S | 1 | 0.4% | 0 (0%) | 0 (0%) | 0 (0%) | 1 (1%) |
| T250I | 1 | 0.4% | 0 (0%) | 0 (0%) | 0 (0%) | 1 (1%) |
| P251L | 2 | 0.9% | 0 (0%) | 0 (0%) | 0 (0%) | 2 (2%) |
| A352S | 1 | 0.4% | 0 (0%) | 1 (1%) | 0 (0%) | 0 (0%) |
| N439K | 1 | 0.4% | 1 (25%) | 0 (0%) | 0 (0%) | 0 (0%) |
| K444N | 1 | 0.4% | 0 (0%) | 1 (1%) | 0 (0%) | 0 (0%) |
| P479S | 1 | 0.4% | 0 (0%) | 0 (0%) | 0 (0%) | 1 (1%) |
| A522S | 1 | 0.4% | 0 (0%) | 0 (0%) | 0 (0%) | 1 (1%) |
| E583D | 1 | 0.4% | 0 (0%) | 1 (1%) | 0 (0%) | 0 (0%) |
| S640F | 1 | 0.4% | 0 (0%) | 1 (1%) | 0 (0%) | 0 (0%) |
| A653V | 1 | 0.4% | 0 (0%) | 1 (1%) | 0 (0%) | 0 (0%) |
| A688V | 1 | 0.4% | 0 (0%) | 1 (1%) | 0 (0%) | 0 (0%) |
| T719I | 3 | 1.3% | 0 (0%) | 0 (0%) | 0 (0%) | 3 (3%) |
| K795R | 1 | 0.4% | 0 (0%) | 1 (1%) | 0 (0%) | 0 (0%) |
| P812L | 1 | 0.4% | 0 (0%) | 1 (1%) | 0 (0%) | 0 (0%) |
| D843N | 1 | 0.4% | 0 (0%) | 1 (1%) | 0 (0%) | 0 (0%) |
| D936H | 1 | 0.4% | 0 (0%) | 0 (0%) | 0 (0%) | 1 (1%) |
| V976F | 1 | 0.4% | 0 (0%) | 0 (0%) | 0 (0%) | 1 (1%) |
| A1056V | 1 | 0.4% | 0 (0%) | 1 (1%) | 0 (0%) | 0 (0%) |
| Q1071L | 2 | 0.9% | 2 (50%) | 0 (0%) | 0 (0%) | 0 (0%) |
| G1085R | 1 | 0.4% | 0 (0%) | 1 (1%) | 0 (0%) | 0 (0%) |
| I1115V | 1 | 0.4% | 0 (0%) | 1 (1%) | 0 (0%) | 0 (0%) |
| S1147L | 1 | 0.4% | 0 (0%) | 1 (1%) | 0 (0%) | 0 (0%) |
| T1160I | 1 | 0.4% | 0 (0%) | 1 (1%) | 0 (0%) | 0 (0%) |
| G1219V | 3 | 1.3% | 0 (0%) | 3 (3%) | 0 (0%) | 0 (0%) |
| V1228L | 1 | 0.4% | 0 (0%) | 1 (1%) | 0 (0%) | 0 (0%) |
| I1232L | 4 | 1.8% | 0 (0%) | 4 (4%) | 0 (0%) | 0 (0%) |
| M1237V | 1 | 0.4% | 0 (0%) | 0 (0%) | 0 (0%) | 1 (1%) |
| V1264L | 2 | 0.9% | 0 (0%) | 1 (1%) | 0 (0%) | 1 (1%) |
| L1265I | 8 | 3.5% | 0 (0%) | 0 (0%) | 0 (0%) | 8 (8%) |

<sup>a</sup>N=225, consisting of 4 non-VOC, 112 Alpha, 3 Gamma, and 106 Delta isolates. <sup>b</sup>Not applicable, this
mutation is a defining mutation of the Gamma variant.

**Figure 3.**
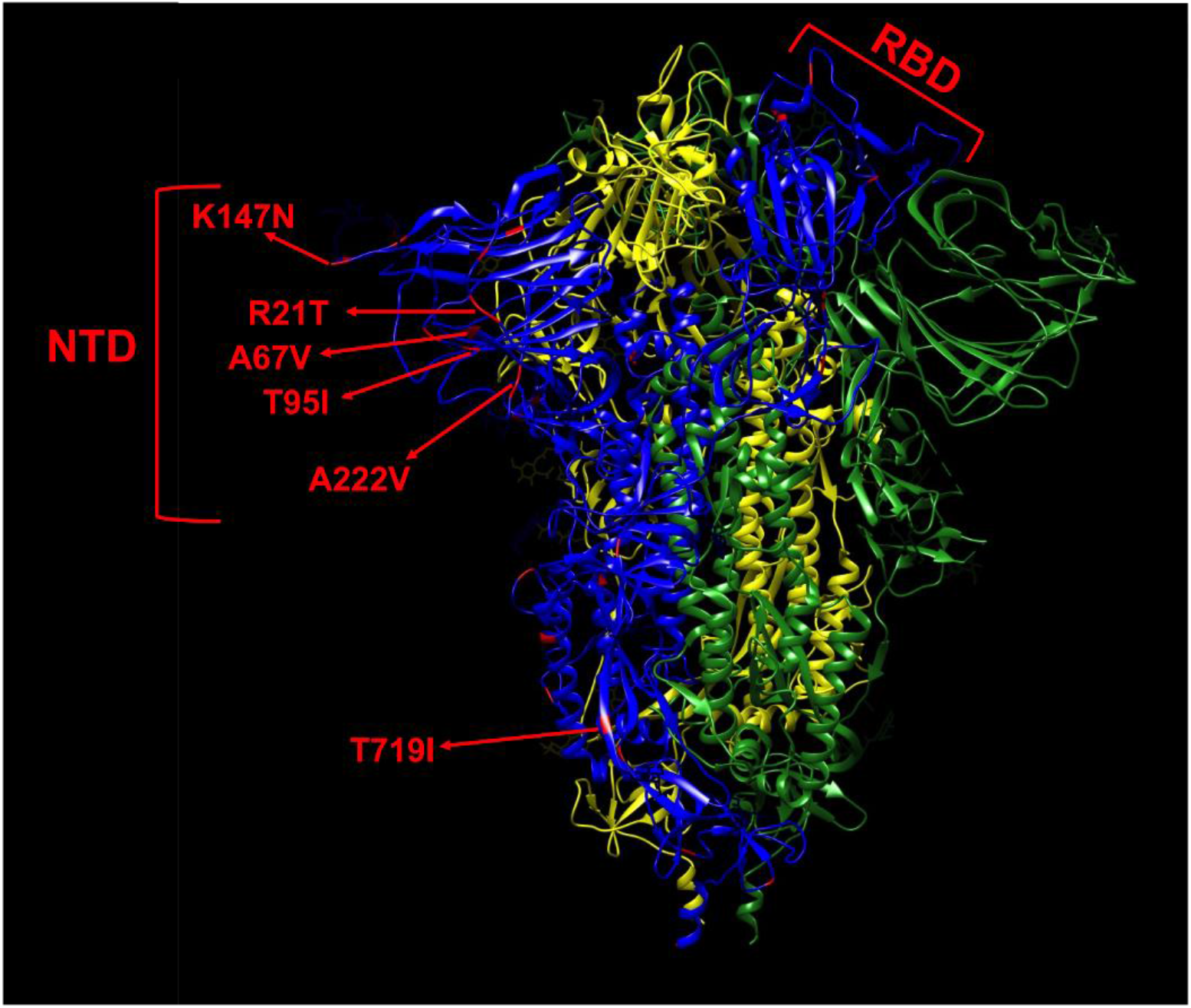
Mapping of non-lineage defining mutations (red) on one monomer (blue) of the spike protein. The most frequent mutations (>1%) are indicated by red arrows. The L5F, G1219V, I1232L, and L1265I mutations are not shown in this model due to their proximity to the termini as the model only comprises residues 14-1147. The spike is shown in its closed state as a trimer of 3 monomers colored in blue, yellow, and green, respectively. NTD, N-terminal domain. RBD, receptor binding domain.

### People experiencing severe symptoms were significantly older than those who remained asymptomatic or experienced mild to moderate symptoms

A significant difference in median age was found between asymptomatic (46.5 years old) and severe symptomatic cases (83 years old) on one hand, and mild-moderate symptomatic (42 years old) and severe COVID-19 cases on the other hand (Figure 4A). Two out of seven severe SARS-CoV-2 infections had a fatal outcome. Nevertheless, the great majority of cases (75.4%) belonged to the mild-moderate category. There was no association between SARS-CoV-2 genotype and disease severity (Table S3).

**Figure 4.**
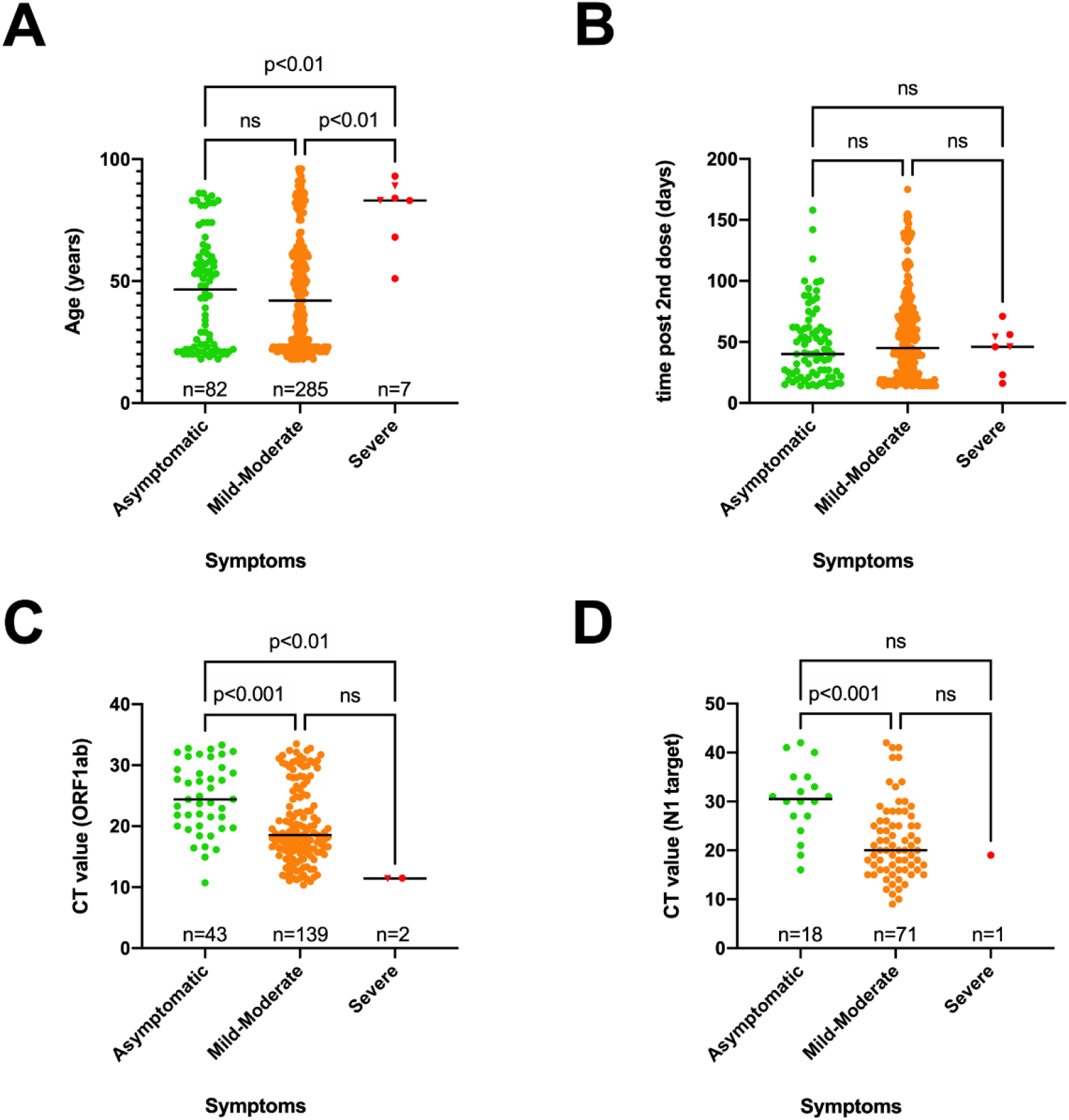
**A**. Scatter plots showing the distribution of age vs symptoms (**A**), time of positive RT-PCR test after receiving the 2^nd^ dose vs symptoms (**B**), CT value vs symptoms for the ORF1ab target (**C**) or N1 target (**D**) as determined using two independent workflows. A Kruskal-Wallis test was performed to compare the median age (**A**), median time of positive RT-PCR test after receiving the 2^nd^ dose (**B**) and the median CT value (**C-D**) between the 3 categories of symptoms. When a statistically significant result was obtained, Dunn’s multiple comparison test was performed *post hoc* to identify which groups significantly differed. ns, not significant. A fatal outcome was indicated by a reverse triangle symbol.

When the time of the first positive test after receiving the 2^nd^ dose was plotted against disease severity for each case, no significant difference could be observed (Figure 4B).

However, a significant difference in median CT value was observed between the three different groups of symptoms, as lower median CT values were observed in the severe and mild-moderate categories compared to asymptomatic cases (Figure 4C&D).

### Relationship between vaccine type and viral load, time of positive test post vaccination and age

No significant difference in viral load could be observed when comparing the infections after Pfizer-BioNTech, Moderna, Oxford-AstraZeneca and J&J/Janssen vaccinations (Figure 5A&B).

**Figure 5.**
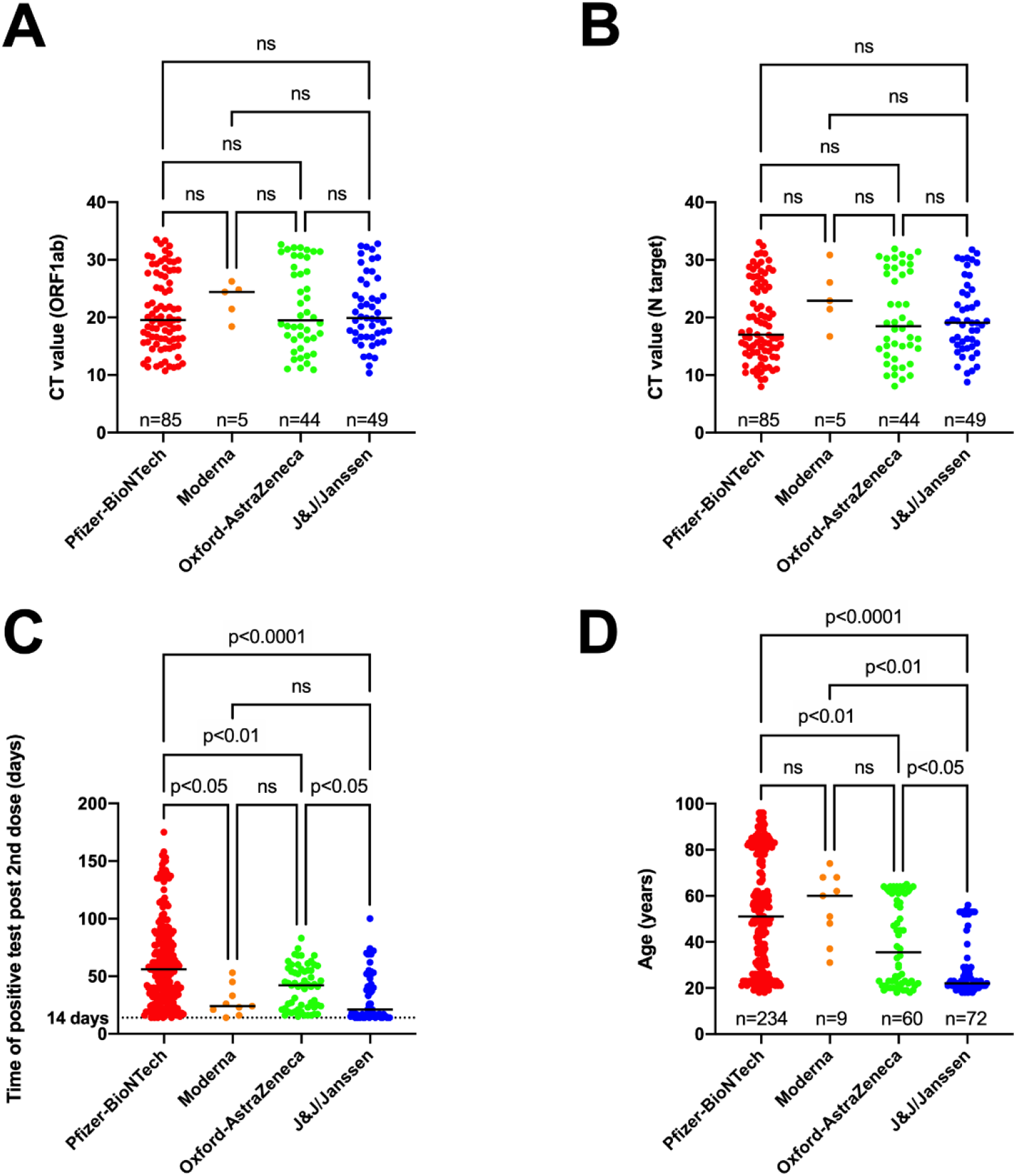
**A**. Relationship between type of vaccine used and CT value obtained for ORF1ab (**A**), N target (**B**), time of positive SARS-CoV-2 test after receiving the 2^nd^ dose (**C**) or age (**D**). A Kruskal-Wallis test was performed to compare median CT value for ORF1ab (**A**), N target (**B**), the median time of positive RT-PCR test after receiving the 2^nd^ dose (**C**) and the median age (**D)** between the 4 types of vaccines. When a statistically significant result was obtained, Dunn’s multiple comparison test was performed *post hoc* to identify which groups significantly differed. ns, not significant.

However, there was a significant difference in median time of the first positive test after receiving the 2^nd^ dose between the different vaccine types as people who received the Pfizer-BioNTech vaccine tested positive later after being fully vaccinated compared to the other vaccine types (Figure 5C). In addition, people who received the J&J/Janssen vaccine in our study received a positive result sooner after being fully vaccinated compared to people receiving the Oxford-AstraZeneca vaccine.

Finally, individuals who received the Pfizer-BioNTech vaccine were significantly older (median age 51 years old) than those who received the Oxford-AstraZeneca (median age 35.5 years old) or the J&J/Janssen vaccine (median age 22 years old) (Figure 5D).

There was no association between vaccine type and disease severity (Table S4).

## Discussion

In this study, we studied breakthrough infections per variant and vaccine type in function of time in the South Limburg region of the Netherlands. We determined that prior to emergence of the Delta variant, breakthrough cases occurred at a continuously low frequency and the proportion of SARS-CoV-2 variants causing breakthrough infections corresponded well with the distribution of those variants in the region. However, both the number of positive cases and breakthrough infections steeply increased starting from week 27, most likely as a consequence of relaxation of COVID-19 measures and subsequent higher virus transmission rates, in The Netherlands by the end of week 25. Furthermore, the proportion of breakthrough cases versus total cases seemed increased. This rise started from week 25, when the Delta variant became dominant in the region. However, whether this increase in relative proportion of breakthrough infections is related to the rise of the Delta variant is difficult to determine since the number of fully vaccinated individuals also quickly accumulated during that period. Other studies have observed a similar impact of the Delta variant as for example in the Delaware valley, the Delta variant showed three-fold enrichment in vaccine breakthrough cases compared to genomic surveillance data through summer 2021^31^. Furthermore, several studies have found that vaccine effectiveness for multiple vaccine types was significantly reduced against the Delta vs Alpha variant^24,32-35^. For instance, a large study assessing breakthrough infections in frontline workers conducted by the CDC revealed that vaccine effectiveness dropped from 91% before predominance of the Delta variant to 66% when this variant became predominant^32^. A second study analyzing breakthrough infections in 780,225 US veterans found that vaccine effectiveness dropped from 87.9% in February when the Delta variant was absent to 48.1% in October when the Delta variant was predominant in the US^35^.

In our study, we also noticed a disproportionate representation of administered vaccine types among breakthrough cases, as the mRNA-based vaccines (0.14%) led to fewer breakthrough infections, compared to the J&J/Janssen (0.46%) or Oxford-AstraZeneca vaccines (0.89%) relative to the number of administered vaccines. Since the number of fully vaccinated people with Oxford-AstraZeneca and J&J/Janssen only significantly started accumulating staring from week 21 and nearly half of the breakthrough cases were recorded between week 25 and week 28, this applies in particular to the Delta variant. A similar observation has been reported by another study in which it was shown that vaccine effectiveness for the J&J/Janssen vaccine dropped from 86% in March to 13% in September, while the decline for the Moderna (89% in March to 58% in September) and Pfizer-BioNTech (87% in March to 43% in August) was much more modest^35^. Furthermore, the Washington DC Health department also found a higher proportion of breakthrough infections among people vaccinated with J&J/Janssen (2.20%) as compared to Pfizer-BioNTech (1.23%) or Moderna (0.86%) relative to the number of complete vaccinations with the respective vaccine types between January and October^36^. Finally, a third study showed that the Oxford-AstraZeneca vaccine was 60% effective against infection with the Delta variant compared to 79% effectiveness for the Pfizer-BioNTech vaccine, despite both vaccine types being 13% less effective compared to infection with the Alpha variant^34^.

When comparing the distribution of SARS-CoV-2 variants retrieved from fully vaccinated individuals compared to the frequency of variants that were circulating in the entire population in the region of South Limburg, we found that breakthrough infections were caused by SARS-CoV-2 genotypes that were present in similar proportions during genomic regional surveillance. Before the Delta variant emerged, the great majority of breakthrough infections were caused by the Alpha variant with the exception of an occasional Gamma or a non-VOC infection. This is in line with data from the CDC in which 555 SARS-CoV-2 isolates from a total of 10,262 SARS-CoV-2 vaccine breakthrough infections were sequenced and the proportion of VOCs identified in breakthrough infections (64%) corresponded with their prevalence in the national genomic surveillance (70%) during the period between January 1 and April 30, 2021^32^.

When the Delta variant emerged in the South Limburg region in week 23, the number of positive cases was low and this variant quickly became dominant in the following weeks when there was a surge in the number of positive cases, making it difficult to estimate what proportion of breakthrough infections are attributable to the Delta variant as compared to regionally surveillance data. Nevertheless, the proportion of breakthrough infections caused by the Delta variant seems to be slightly higher than its share in genomic surveillance (100% vs 64% in week 26, 95% vs 84% in week 27, and 100% vs 95% in week 28), although still being in line with our observations for the Alpha variant and other VOCs.

Marques and colleagues^31^ found a much more pronounced enrichment of the Delta variant (3-fold) in SARS-CoV-2 breakthrough infections compared to its frequency in surveillance data in the Delaware valley. The fact that the time window from emergence to predominance of the Delta variant was wider (2 months vs 1 month) in the Delaware study might explain this discrepancy.

When looking at the frequency of non-lineage defining mutations in the 225 isolates causing breakthrough infections in this study, it was found that the great majority of mutations were concentrated in the N-terminal and C-terminal domains of the spike protein. Amongst the 3 most frequent non-lineage defining mutations were the T95I (4.9%), R21T (10.4%), and A222V (28.4%) amino acid substitutions in the NTD, of which the latter two were detected in different variant backgrounds. It is known that mutations in the antigenic supersite of the N-terminal domain (NTD) of the spike protein (residues 14-20, 140-158, and 245-264) can lead to reduced neutralization by a number of monoclonal antibodies^16^ For instance, mutation of residues 156-158 have been described in B.1.1.523^37^, being a variant under monitoring (VUM) and the Delta variant^25^. Although the A222V and T95I mutations are in close proximity to the antigenic supersite residues, they have no effect on neutralization^16^. Nevertheless, the effect of these mutation in the Delta background on neutralization has not been determined yet and the T95I mutations is found in many VUM/VOI, including the B.1.1.318, Kappa, Iota, and Mu variants^5,38^ and as well in an emerging sublineage of the Delta variant^39^. Furthermore, the A222V mutation has emerged independently in the B.1.177 and two Delta sublineages, including the AY4.2 (“Delta Plus”) lineage that is rapidly increasing in frequency at the moment in the UK^39^. In addition, all A222V mutations in this study belonged to the AY.9 sublineage of the Delta variant that has also been associated with breakthrough infections in India^40^.

The finding that the majority of adaptive mutations are concentrated in the NTD corresponds well with recent data published by Zhang *et al*, showing that different variants tolerate multiple mutations causing structural rearrangements in their NTD that lead to immune evasion, while the overall structure of the RBD is strictly preserved, with mutations only being limited to a number of sites^41^.

Previously, we have shown that infections caused by the Delta variant are associated with higher viral loads compared to the Alpha variant in the total population^42^. Here, we found that this is also the case for a fully vaccinated population as a median difference of 5 CT-values was observed between infections caused by the Delta vs Alpha variant. A similar difference in load between both variants was observed in the study of Blanquart *et al*, in which the CT values of 77 non-Delta (mostly Alpha) breakthrough infections were compared to 866 breakthrough cases infected with the Delta variant^43^. A large study including 16,000 infected individuals during the Delta-dominant period in Israel found that fully vaccinated individuals harbored lower viral loads than unvaccinated individuals, but that the effect of vaccination on viral loads started declining after 2 months and completely vanished 6 months post vaccination^44^. However, the difference was restored after a booster dose of the Pfizer-BioNTech vaccine. This indicates that viral loads found in vaccinated and unvaccinated individuals might be comparable in a relatively short amount of time post vaccination.

When looking at the distribution of disease symptoms in our study population, a significant difference in median age was observed between asymptomatic and severe (46.5 vs 83 years) as well as mild-moderate and severe cases (42 vs 83 years). Similar findings were reported in Belgium^45^ and the USA^46^, where the median age of hospitalized patients was 82 and 80.5 years, respectively.

A number of hypotheses could explain the breakthrough infections causing mortality in the oldest age group. First of all, underlying conditions could have increased the vulnerability to SARS-CoV-2 infections in these individuals. Secondly, the humoral and cellular immune response triggered by the vaccines might not have been adequate to protect from symptomatic SARS-CoV-2 infection. Recently, it has been shown that there was a significant correlation between increased age and reduced antibody neutralization when sera from individuals vaccinated with the BNT162b2 vaccine of Pfizer were incubated with either wild-type, D614G, B.1.1.7, B1.351 or B.1.617.2 SARS-CoV-2 virus^20^. Furthermore, a second study reported that a weaker T cell response was observed in older individuals following vaccination^47^. It has been proposed that age-related thymic involution causes immunosenescence due to reduced T cell receptor (TCR) diversity as well as increased chronic inflammation in the elderly^48^. Reduced TCR diversity could interfere with the ability to respond to novel antigens, while increased chronic inflammation could induce a cytokine storm^48^.

No significant difference in time of positive test after the 2^nd^ dose could be observed between asymptomatic patients, patients with mild-moderate or severe symptoms in our study. However, it has been described that vaccine efficacy can drop over time as much as 22% for Pfizer-BioNTech and 7% for Oxford-AstraZeneca when comparing 14 days to 90 days post receiving the 2^nd^ dose^33^.

Interestingly, we found that symptomatic patients harbored significantly higher viral loads compared to asymptomatic people amongst breakthrough cases. A similar observation has been done by Shamier and colleagues in The Netherlands^49^ and by Blancqaert *et al* in France^43^.

In agreement with the study of Shamier et al^49^, no significant difference in viral load could be found when comparing the different types of vaccines that had been administered, despite the median CT-value for the few (n=5) infections in Moderna recipients being 5 units higher. A recent study has shown that higher neutralizing antibodies within a week before SARS-CoV-2 infection are associated with lower viral loads^50^. It would be interesting to see if the Moderna vaccine is associated with lower viral loads compared to other vaccine types in a larger study of breakthrough infections, as higher antibody titers have been observed in people receiving 2 doses of Moderna compared to 2 doses of Pfizer-BioNTech^51^, most likely due to the >3-fold higher spike mRNA content.

The significant increase in age and time of the first positive test after receiving the 2^nd^ dose observed for the Pfizer-BioNTech compared to the Oxford-AstraZeneca and J&J/Janssen vaccines can be explained by the fact that during the first 21 weeks mainly Pfizer-BioNTech was used to inject mainly the elderly and healthcare workers, whereas the J&J/Janssen vaccine was administered to young adults who received their second dose right before relaxation of COVID-19 measures when there was a spike in positive cases.

Strengths of this study were the ability to monitor and compare breakthrough infections against the regional genomic surveillance over an extensive period, even before the first vaccines were administered as well as the regional follow-up of cases, allowing monitoring of the amount of breakthrough infections at a high resolution. A high proportion (60%) of all breakthrough cases were successfully sequenced further enhancing the resolution. Additionally, analysis were conducted on cases one day after vaccination giving an overall picture of susceptibility directly post vaccination.

Limitations were the fact that the number of reported breakthrough infections are an underestimation of the effective number, due to asymptomatic or mild cases or because of severe cases in local hospitals that were not reported to the regional public health service. Furthermore, asymptomatic cases were only followed up for a limited amount of time, possibly leading to the underestimation of symptomatic cases. Thirdly, only samples from cases with sufficiently high viral loads could be sequenced (<CT 32), leading to a limited dataset. Additionally, not all samples were available so some bias could have been introduced if unavailable samples were different than available samples. Lastly, the lack of immunological data, which could help to understand how breakthrough infections occur and why certain individuals experience severe symptoms.

In summary, this study has investigated the distribution of breakthrough infections per vaccine type and variant over time in the South Limburg region of The Netherlands. It was found that breakthrough infections were more frequently observed in people who received Oxford-AstraZeneca or J&J/Janssen in comparison with recipients of the mRNA-based vaccines. Furthermore, the predominance of the Delta variant coincided with a rapid increase in breakthrough infections and severe cases were only observed in older individuals. Given that reduced vaccine effectiveness for the Delta variant has been reported by multiple studies, as well as the fact that the Delta variant is associated with increased viral loads in both vaccinated and unvaccinated populations might suggest that a booster shot for certain vaccine types and populations at risk might be helpful in precluding future COVID-19 waves.

## Data Availability

All data produced in the present study are available upon reasonable request to the authors. SARS-CoV-2 whole genome sequences were deposited in the GISAID database.

## Data availability

All sequences have been uploaded to GISAID. All GISIAD accession ID numbers can be found in Table S2.

## Acknowledgements

We would like to thank Edou Heddema from Zuyderland MC and Boris Vlaemynck from Synlab Belgium for providing samples and data.

## Supplementary material

**Table S1.**
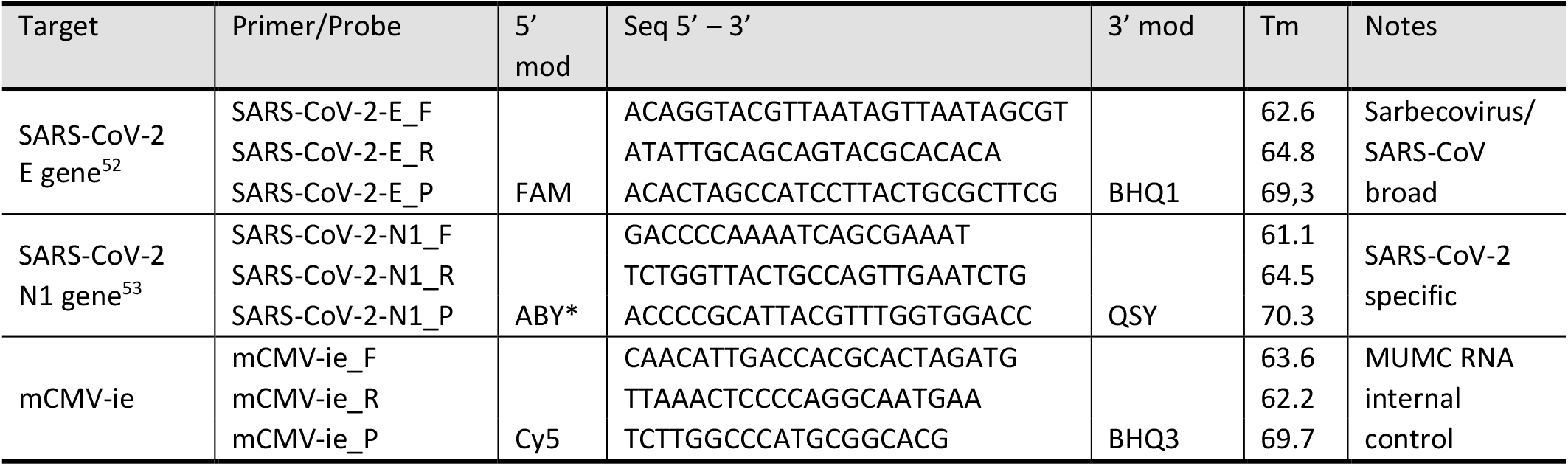
Oligonucleotides used in this study.

**Table S2.** Cohort metadata.

**Table S3.** Contingency table of genotype vs symptoms

| N (Percentage of grand total) | Asymptomatic | Mild-Moderate | p-value* |
| --- | --- | --- | --- |
| Alpha | 22 (10.19%) | 88 (40.74%) | 0.28 |
| Delta | 15 (6.94%) | 91 (42.13%) |  |

\*p-value was calculated using Fisher's exact test.

**Table S4.** Contingency table of vaccine type vs symptoms

| N (Percentage of grand total) | Asymptomatic | Mild-Moderate | p-value* |
| --- | --- | --- | --- |
| Pfizer-BioNTech | 53 (14.25%) | 178 (47.85%) | 0.96 |
| Moderna | 2 (0.54%) | 7 (1.88%) |  |
| Oxford-AstraZeneca | 12 (3.23%) | 48 (12.90%) |  |
| J&J/Janssen | 15 (4.03%) | 57 (15.32%) |  |

\*p-value was calculated using a Chi-square test.

